# Cardiovascular, renal and pulmonary risks of Long COVID: a retrospective cohort study stratified by age and sex

**DOI:** 10.64898/2026.01.14.26344156

**Authors:** Akash Pradeep, Martha L Carvour, Srija M Sivamurugan, Upinder Singh, Alejandro P Comellas, Sanjana Dayal

**Affiliations:** Department of Internal Medicine, University of Iowa Carver College of Medicine, Iowa City, Iowa; Fraternal Order of Eagles Diabetes Research Center, University of Iowa Carver College of Medicine; Abboud Cardiovascular Research Center, University of Iowa Carver College of Medicine, Iowa City, Iowa; Holden Comprehensive Cancer Center, University of Iowa Carver College of Medicine, Iowa City, Iowa; Iowa City VA Healthcare System, Iowa City, Iowa

## Abstract

**Background:** A substantial proportion of patients with acute COVID-19 develop post-acute sequelae of SARS-CoV-2 infection (Long COVID). The risk of adverse cardiovascular and related outcomes in Long COVID remain elusive. We hypothesized that individuals with Long COVID are at elevated risk for adverse cardiovascular, renal and pulmonary (CRP) outcomes compared to those who recovered from COVID-19 without developing Long COVID.

**Methods:** We performed a retrospective cohort study using the global TriNetX electronic health record network (> 150 million patients). Adults with documented COVID-19 were classified by the presence/absence of Long COVID. We analyzed absolute risks (AR) and relative risks (RR) for 15 CRP outcomes, stratified by age (18–50, 51–64, ≥65 years). After excluding those with the preexisting outcomes of interest, propensity score matching was applied to adjust for age, sex, and common confounders.

**Results:** Among 2,613,432 adults with COVID-19, 315,612 matched individuals were included in the study. Long COVID was associated with increased AR for most CRP outcomes across adult ages regardless of sex. RR were disproportionately higher in younger adults, especially in females for cardiovascular and renal outcomes and in males for selected pulmonary outcomes. In a secondary analysis among individuals with Long COVID, prior COVID-19 vaccination was not associated with a significantly lower risk of CRP outcomes.

**Conclusions:** Long COVID is associated with increased risk of adverse CRP outcomes, with relatively higher risks observed in younger adults. These findings support the need for continuing surveillance and risk-reduction strategies for cardiovascular and related disorders in Long COVID.

## Introduction

With the increasing use of vaccines and advancement of effective medications, the threat of COVID-19 due to SARS-CoV-2 infection has decreased.^1–3^ However, attention has shifted from managing acute infections to addressing the long-term consequences of infection, particularly post-acute sequelae of COVID-19 (Long COVID or post-COVID conditions).^4^ Long COVID is estimated to affect 10-25% patients with COVID-19 worldwide.^5^ Several studies have reported elevated long-term risk for new-onset cardiovascular, renal and pulmonary (CRP) complications as a consequence of COVID-19.^6–10^ However, it is unclear whether these risks in patients with Long COVID exceed those observed in individuals who recover from acute COVID-19 without developing Long COVID.

Existing studies are inadequate to address this question, with most conducted within the first two years of the pandemic and did not distinguish cohorts based on Long COVID status.^6–10^ These reports are further limited by either modest sample sizes, regional cohorts, narrow outcome definitions, or inadequate matching for baseline comorbidities. Moreover, stratification of outcomes by age or implications of sex differences within each age strata have not been evaluated, despite the evidence that these factors modulate chronic COVID-19 outcomes. As SARS-CoV-2 continues to evolve, refining our understanding of these enduring long-term multi-organ health consequences of infection across diverse populations, and identifying age-and sex-specific vulnerabilities, is critical for informing need for mechanistic studies, post-acute care, and surveillance strategies.

In this retrospective cohort study, we tested the hypothesis that individuals with Long COVID are at higher risk for CRP complications compared to individuals who recovered from COVID-19 without developing Long COVID. Leveraging the TriNetX global federated electronic health record network, we conducted a large-scale rigorous propensity score matched retrospective cohort analysis to assess age- and sex-specific risks of CRP outcomes independent of baseline comorbidities/confounders. Secondly, we also assessed whether prior vaccination modifies the risks of CRP outcomes in Long COVID. Our findings provide a framework for age-and sex-specific risk stratification to guide post-acute care planning for cardiovascular and related outcomes, and with implications to mitigate the chronic burden of Long COVID.

## Methods

### Setting and Study Populations

In this retrospective cohort study, we acquired data from the TriNetX, a global federated health research network with real-time updates of anonymized electronic health records (EHRs) provided by health care organizations (HCOs) worldwide. Patients were selected from TriNetX using International Classification of Diseases version 10 (ICD-10) codes documented between June 2020 through December 2024. We included two adult groups (age ≥18 years) for comparison: individuals with post-COVID condition (Long COVID, ICD-10 code: U09) and individuals with a diagnosis for COVID-19 without a diagnosis for post-COVID condition (control cohort, identified using a TriNetX-curated code for positive SARS-CoV-2 test). The index event for the Long COVID cohort was defined as the time of the first diagnosis of post-COVID condition. The patients in control cohorts were matched by age and sex with the Long COVID patients at the time of their interaction with healthcare. Both cohorts were stratified into age groups defined as young (18-50 years), middle-aged (51-64 years), and senior (65 and higher [≥ 65]) adults for analysis.

### Ethics statement

The TriNetX platform is compliant with the Health Insurance Portability & Accountability Act and General Data Protection Regulation. The Western Institutional Review Board has granted TriNetX a waiver of informed consent since this platform uses only aggregated counts and statistical summaries of de-identified information, therefore, this study is exempt from informed consent. The data reviewed is a secondary analysis of existing data, does not involve intervention or interaction with human subjects, and is de-identified.

### Assessment of Pre-specified Outcomes, Matching and Analytics

Outcomes were compared across the matched cohorts with and without post-COVID condition using four separate models representing the following outcomes (**Figure 1)**:

*a) Cardiovascular outcomes*: essential hypertension (HTN [I10]), atrial fibrillation (I48.91), ischemic heart diseases (IHD, I20-I25), and heart failure (HF, I50).
*b) Thrombotic outcomes*: deep vein thrombosis (DVT, I82.40), pulmonary embolism (PE, I26), acute myocardial infarction (MI, I21), and cerebral infarction or ischemic stroke (IS [I63]).
*c) Renal outcomes*: acute kidney injury (AKI, N17), chronic kidney disease (CKD, N18), and end stage renal disease (ESRD, N18.6).
*d) Pulmonary outcomes*: pulmonary fibrosis (PF, J84.10), pulmonary hypertension (PHT, I27.20), chronic bronchitis (CB, J42) and chronic respiratory failure (CRF, J96.1).

**Figure 1:**
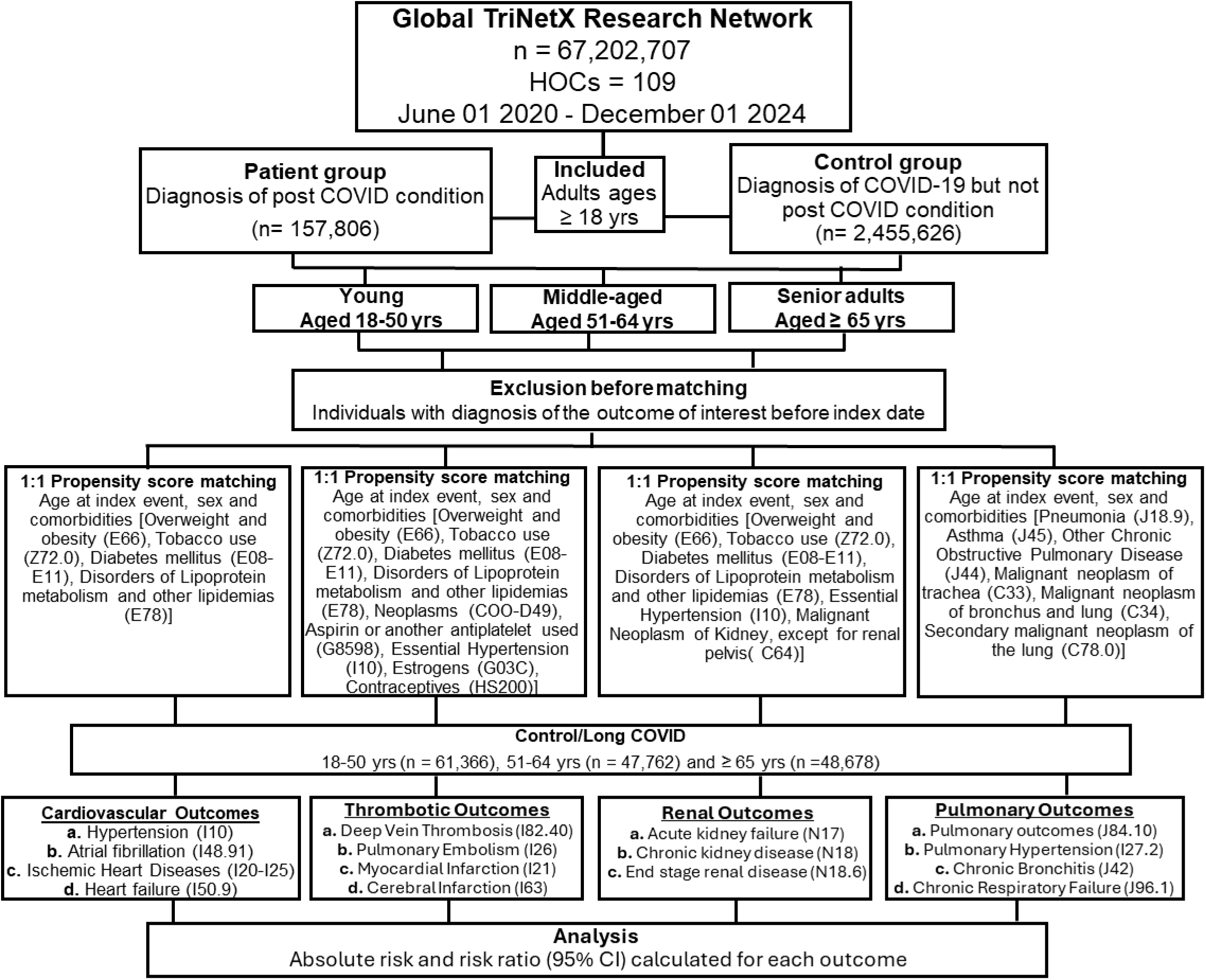
Flow Chart of Study design.

For each outcome analysis, 1:1 propensity score matching was performed within each age group to match the groups by the age, sex and confounders (comorbidities and medications that are known to affect outcome of interests). Covariates used in the analysis, and confounders included in the matching process, were defined based on previous knowledge from clinical practice and salient literature. The confounders that were used for matching for cardiovascular outcomes included overweight and obesity (ICD-10 code; E66), diabetes mellitus (E08-513), dyslipidemia (disorders of lipoprotein metabolism and other lipidemia [E78]), or use of tobacco (C78.0). In addition to these confounders, we also matched for HTN, neoplasms (C00-D49), aspirin or another antiplatelets (G8598), estrogens (G03C) or systemic contraceptives (HS200) for analyzing thrombotic outcomes. For renal outcomes, we matched for tobacco use, HTN, diabetes mellitus, dyslipidemias and malignant neoplasms of kidney (C64). Finally, for pulmonary outcomes, we matched for pneumonia (J18), asthma (J45), chronic obstructive pulmonary disease (COPD [J44]), tobacco use, secondary malignant neoplasm of trachea (C33), bronchus and lung (C34).

To exclude outcomes already present at baseline, including those present at the time of a Long COVID diagnosis, we excluded all events meeting these outcome criteria that occurred prior to the diagnosis of Long COVID. Likewise, to exclude conditions already present at baseline or occurring during the counterfactual risk period for post-COVID in the control group, we only included events that occurred 2 months after the diagnosis of COVID-19 among controls.

### Vaccination

To assess effects of vaccination, a sub-analysis was performed within the Long COVID cohort where outcomes were compared between those who were vaccinated (SARS-CoV-2 [COVID-19] vaccine, mRNA spike protein, RxNorm code 2468231), or not vaccinated before developing Long COVID. The patient group was defined as those receiving the vaccine before diagnosis of post-COVID, while the control group only had post-COVID condition.

### Statistical Analysis

To limit the influence of confounding factors, we used TriNetX built-in function for propensity score matching and generated groups with matched baseline characteristics. Standardized difference (Std diff) was used to evaluate the balance of baseline characteristics in the propensity score-matched groups. Balance on covariates was assessed using standardized mean difference and values > 0.1 were considered positive for residual imbalance.^11–13^ The absolute risk (AR) and risk ratio (RR) in each age group and within each sex were assessed. The TriNetX calculates RR and associated 95% confidence intervals (95% CI), using R’s Survival package, version 3.2-3 (R Group for Statistical Computing). Chi-square tests and T test were used for univariate analysis, and performed using the built-in statistical tool on the TriNetX platform based on Java, version 11.0.16 (Oracle) with the outputs validated using independent, industry-standard methods.^14^ Statistical significance was defined as P-value < 0.05. The results were provided to investigators in summarized format. Details regarding the database can be found online.^15^

## Results

### Demographic stratification

We identified 2,613,432 adults with COVID-19 in the TriNetX network between June 2020 and December 2024, including 157,806 (6.0%) individuals with post-COVID condition (Long COVID cohort) and 2,455,626 (94.0%) without a subsequent post-COVID diagnosis (control cohort). The demography of both cohorts before and after matching 1:1 with Long COVID patients by age, sex and baseline comorbidities/confounders, and stratified into age groups as young (18-50 years), middle-aged (51-64 years) and senior (≥ 65 years) adults are presented in **supplementary Table S1-S3.** After propensity score matching, the final analytic cohort comprised 315,612 individuals, with 157,806 in each group. Within matched cohorts, there were 61,366 young, 47,762 middle-aged and 28,767 senior adults in each cohort, and nearly three times as many female patients as males in the young and middle-aged groups, and twice as many females in the senior adult group.

Before matching, comorbidities including HTN, overweight and obesity, hyperlipidemia, diabetes, asthma, COPD, pneumonia, and neoplasms were more frequent in Long COVID patients. Propensity score matching balanced these differences, except that the residual higher rates of asthma, COPD, and pneumonia in certain age groups remained. The use of tobacco, aspirin, estrogen and contraceptives were also higher in Long COVID compared to control cohort in most groups and were normalized after propensity score matching. Since ethnicity and race data are incomplete in TriNetX with a large number of unknown values, controlling for them can inadvertently bias causal inferences in observational analyses,^16^ therefore, ethnicity and race were not used as the criteria for matching. However, when propensity score matching was applied for age, sex and comorbidities/confounders, it also normalized the differences for all ethnicities and races in young and middle-aged and for most of these in the senior adults **(Supplementary Table S4-S6**).

So, overall, our cohort was adequately balanced and provided a robust foundation to assess age- and sex-specific absolute and relative risks for CRP outcomes.

### Long COVID patients exhibit an increased risk of several cardiac outcomes

In this propensity score matched analysis, the Long COVID compared to control cohort was found associated with significant increase in risk for several major cardiovascular complications across all age groups, with an expected age-dependent increase seen in both cohorts **(Figure 2).** For HTN, the risk was higher in young adults (AR 6.1% vs 5.0% [RR 1.21, 95% CI 1.15, 1.27]), middle-aged (AR 11.5% vs 9.8% [RR 1.17, 95% CI 1.11, 1.23]), and seniors (AR 16.4% vs 14.3% [RR 1.14, 95% CI 1.12, 1.25]) with Long COVID **(Figure 2A)**. For AF, young adults with Long COVID had nearly twice the risk (AR 0.45% vs 0.26% [RR 1.73, 95% CI 1.43, 2.09]), and elevated risks were also evident in middle-aged (AR 1.71% vs 1.35% [RR 1.27, 95% CI 1.15, 1.41]) and seniors (AR 4.64% vs 4.30% [RR 1.08, 95% CI 1.01, 1.15]) (**Figure 2D)**. For IHD, relative risk was highest in young adults (AR 1.92% vs 1.10% [RR 1.74, 95% CI 1.59, 1.91]) and was sustained in middle-aged (AR 6.77% vs 4.55 [RR 1.49, 95% CI 1.41, 1.57]) and senior adults (AR 11.38% vs 8.37% [RR 1.36, 95% CI 1.30, 1.42]) **(Figure 2G)**. For HF also, Long COVID patients had consistently higher risks, most pronounced in young adults (AR 1.05% vs 0.60% [RR 1.75, 95% CI 1.54, 1.99]), followed by middle-aged (AR 3.16% vs 2.23% [RR 1.42, 95% CI 1.31, 1.53]) and senior adults (AR 7.01% vs 5.83% [RR 1.20, 95% CI 1.14, 1.27]) **(Figure 2H)**.

**Figure 2:**
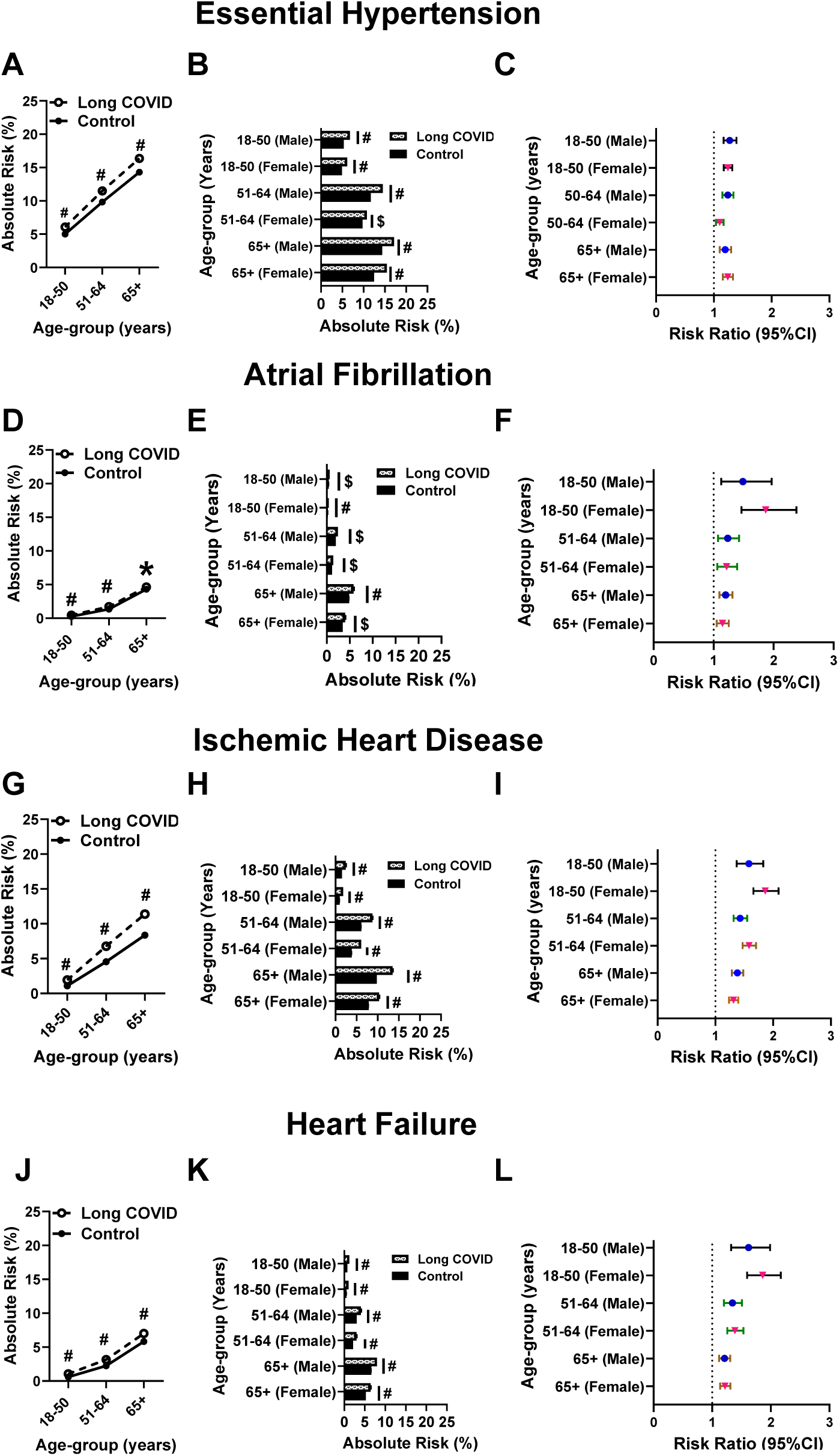
Cardiovascular outcomes in Long COVID. Age and sex based absolute risk, and risk ratios of essential hypertension **(A-C)**, atrial fibrillation **(D-F)**, ischemic heart disease **(G-I)**, and heart failure **(J-L)** in Long COVID. *P<0.05, ^$^P<0.01, ^#^P<0.0001. P values refer to chi-squared analysis of risk differences between Long COVID and control.

Sex-stratified analyses showed that despite the greater number of female patients in the Long COVID cohort, the increased risks for all outcomes examined were present in both males and females across age groups (**Figure 2B, 2E, 2H, 2K).** Finally, the RR was highest in the youngest group for all outcomes where females showed a higher increase than males for AF, IHD and HF (**Figure 2C, 2F, 2I and 2L)**.

### The risks of venous thrombosis and cerebral infarction but not MI persist in Long COVID

We next assessed risks for venous and arterial thrombosis. For DVT, AR was significantly higher in patients with Long COVID than in matched controls across all age groups (AR 0·42% vs 0·22% [RR 1·91, 95% CI 1·49, 2·23] in the youngest group; AR 0·82% vs 0·54% [RR 1·52, 95% CI 1·30, 1·78] in middle-aged, and AR 1·18% vs 0·97% [RR 1·21, 95% CI 1·07, 1·37] in seniors) **(Figure 3A)**. Similarly, risk for PE was increased significantly in long COVID across ages (AR 0·53% vs 0·29% [RR 1·80, 95% CI 1·49, 2·14] in the youngest group, AR 0·95% vs 0·64% [RR 1·48, 95% CI 1·28, 1·70] in middle-aged, and AR 1·39% vs 1·09% [RR 1·28, 95% CI 1·14, 1·42] in seniors) **(Figure 3D)**. In contrast, risk for MI did not differ between Long COVID and controls: ARs were 0·45% vs 0·39% [RR 1·15, 95% CI 0·97–1·36] in the youngest group, 1·40% vs 1·32% [RR 1·06, 95% CI 0·95, 1·18] in middle-aged, and 3·41% vs 3·16% [RR 1·08, 95% CI 0·97, 1·36] in seniors **(Figure 3G)**. For stroke, risk was significantly increased in the youngest group (AR 1·04% vs 0·60% [RR 1·48, 95% CI 1·21, 1·75]) and in seniors (AR 2·78% vs 2·43% [RR 1·14, 95% CI 1·01, 1·25]), but not in the middle-aged (AR 1·137% vs 1·032% [RR 1·10, 95% CI 0·95, 1·21]) **(Figure 3J)**.

**Figure 3:**
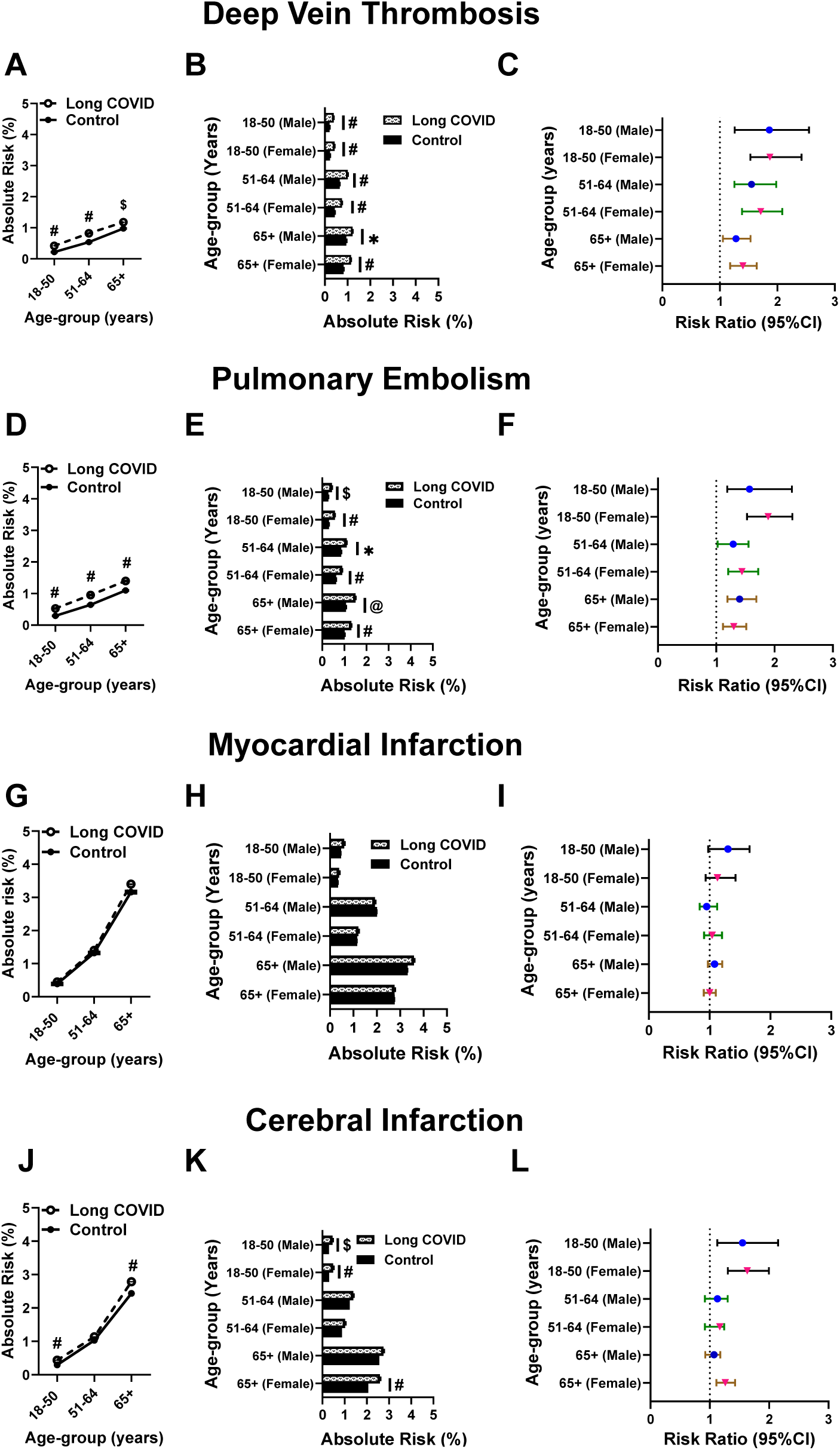
Thrombotic outcomes in Long COVID. Age and sex based absolute risk, and risk ratios of deep vein thrombosis **(A-C)**, pulmonary embolism **(D-F)**, acute myocardial infarction **(G-I)**, and stroke **(J-L)** in Long COVID. *P<0.05, ^$^ P<0.01, ^@^P<0.001, and ^#^P<0.0001. P values refer to chi-squared analysis of risk differences between Long COVID and control.

Sex-stratified analyses showed consistent increases in venous thrombosis across both sexes and all age groups **(Figure 3B, 3E**), but not for MI **(Figure 3H)**. For stroke, increased AR was evident in both sexes in the youngest group, and in females among senior adults **(Figure 3K)**. Across outcomes, RRs were highest in the youngest group, with the largest increase in RR was observed for PE in young females **(Figure 3C, 3F, 3I and 3L).**

### Long COVID is associated with chronic kidney disease in all age groups

We next assessed risk for renal outcomes. For AKI, risk was modestly increased in young (AR 1·15% vs 1·03% [RR 1·11, 95% CI 1·0, 1·24]) and middle-aged (AR 2·92% vs 2·62% [RR 1·12, 95% CI 1·03, 1·20]) Long COVID patients, but decreased in senior adults (AR 6·11% vs 6·64% [RR 0·92, 95% CI 0·87, 0·97]) (**Figure 4A)**. For CKD, Long COVID was consistently associated with higher risk in the young (AR 1·06% vs 0·63% [RR 1·69, 95% CI 1·49, 1·91]), middle-aged (AR 3·29% vs 2·49% [RR 1·32, 95% CI 1·23, 1·43]) and in seniors (AR 7·71% vs 7·05% [RR 1·09, 95% CI 1·04, 1·15]) **(Figure 4D)**. For ESRD, no significant differences were observed between Long COVID and controls: ARs were 0·19% vs 0·17% [RR 1·10, 95% CI 0·85, 1·41] in the young, 0·46% vs 0·52% [RR 0·89, 95% CI 0·75, 1·06] in middle-aged, and 0·79% versus 0·70% [1·14, 95% CI 0·99, 1·31] in seniors **(Figure 4G)**.

**Figure 4:**
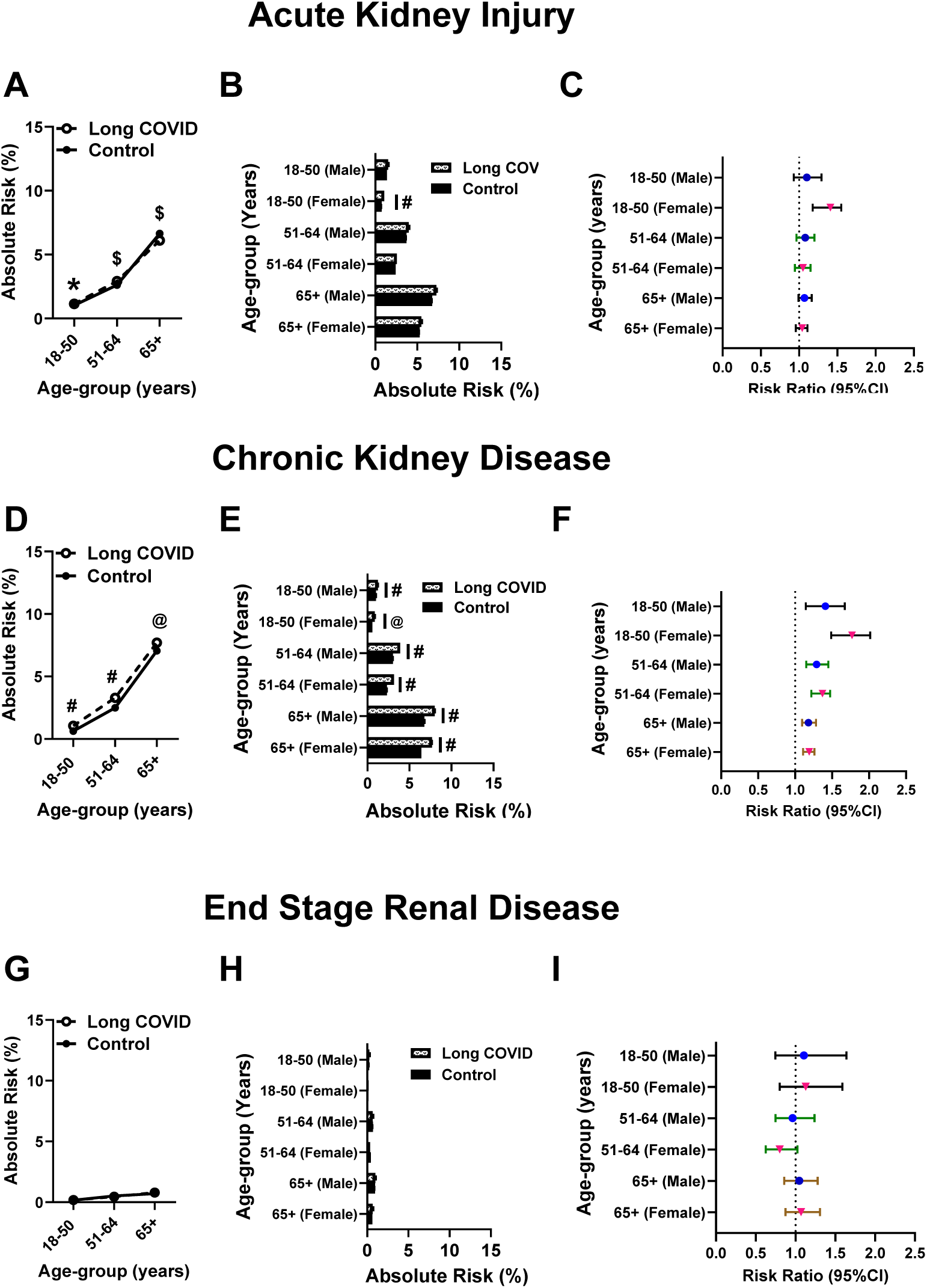
Renal outcomes in Long COVID. Age and sex based absolute risk, and risk ratios of acute kidney injury **(A-C)**, chronic kidney disease **(D-F)** and end satge renal disease (G-I) in Long COVID. *P<0.05, ^$^P<0.01, ^@^P<0.001, and ^#^P<0.0001. P values refer to chi-squared analysis of risk differences between Long COVID and control.

In sex-stratified analyses, a significant increase in AKI was observed only in young females with Long COVID (**Figure 4B)**, whereas CKD risk was significantly higher in both sexes across all age groups (**Figure 4E**). Risk for ESRD did not differ by sex (**Figure 4H**). The greatest increases in RR for both AKI and CKD were observed in the youngest cohort **(Figure 4C, 4F)**, with young females showing higher RRs than males, and RR for ESRD for both sexes remains insignificant (**Figure 4I**).

### Pulmonary disorders continue to persist in Long COVID

Finally, we assessed the risks of pulmonary outcomes in Long COVID compared to the control cohort. For PF, risk was significantly increased across all age groups: AR was 0·54% vs 0·09% [RR 5·58, 95% CI 4·29, 7·25] in the young, AR 1·64% vs 0·34% [RR 4·78, 95% CI 4·06, 5·63] in middle-aged, and AR 2·48% vs 0·96% [RR 2·58, 95% CI 2·33, 2·87] in seniors **(Figure 5A).** For PH, in the youngest group, AR was 0·65% vs 0·26% [RR 2·52, 95% CI 2·11, 3·02], in middle-aged AR was 1·76% vs 0·85% [RR 2·07, 95% CI 1·85, 2·32], and in seniors AR was 3·29% vs 2·65% [RR 1·25, 95% CI 1·16, 1·34] (**Figure 5D)**. For CB, AR was 0·23% vs 0·12% [RR 1·92, 95% CI 1·47, 2·51] in the young, AR 0·71% vs 0·46% [RR 1·56, 95% CI 1·33, 1·83] in middle-aged, and AR 1·09% vs 0·72% [RR 1·50, 95% CI 1·32, 1·71] in seniors (**Figure 5G)**. Likewise, for CRF risk was increased in all age groups: AR was 0·68% vs 0·19% [RR 3·69, 95% CI 3·03, 4·50] in the young, 2·11% vs 1·04% [RR 2·04, 95% CI 1·84, 2·27] in middle-aged, and 3·31% vs 1·91% [RR 1·73, 95% CI 1·60, 1·88] in senior adults **(Figure 5J**).

**Figure 5:**
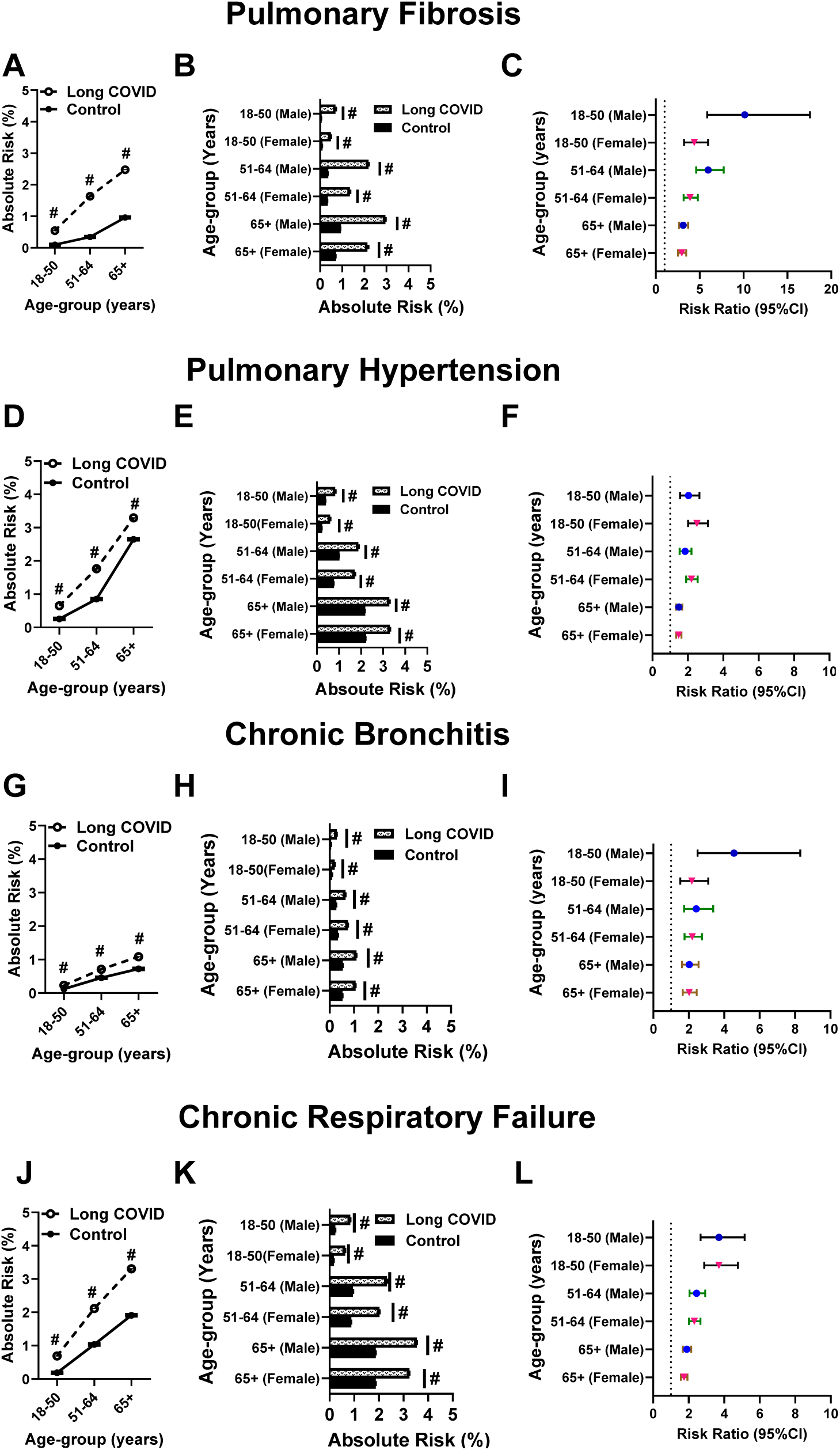
Pulmonary outcomes in Long COVID. Age and sex based absolute risk, and risk ratios of pulmonary fibrosis **(A-C)**, pulmonary hypertension **(D-F)**, chronic bronchitis **(G-I)**, and chronic respiratory failure **(J-L)** in Long COVID. ^#^P<0.0001. P values refer to chi-squared analysis of risk differences between Long COVID and control.

In sex-stratified analyses, significant increases in risk were observed for all pulmonary outcomes in both sexes compared with controls across all age groups **(Figure 5B, 5E, 5H and 5K)**. The greatest increase in RRs was seen for PF, with a 3–10-fold rise across age groups and the highest RR was seen in young males **(Figure 5C)**. For other pulmonary outcomes, RRs ranged from 2 to 5, with the largest increases observed in the youngest cohorts, particularly in young males for CB **(Figure 5F, 5I and 5L)**.

### Vaccination to SARS-CoV-2 did not significantly lower risk for adverse CRP outcomes in Long COVID patients

In secondary analyses, SARS-CoV-2 vaccination prior to Long COVID was not associated with reduced risk of most outcomes. Exceptions included a lower risk of HF in the youngest group (AR 1·13% vs 1·49% [RR 0·76, 95% CI 0·62, 0·93]) and of CRF across all age groups: ARs were 0·65% vs 1·032% [RR 0·63, 95% CI 0·49, 0·82] in the youngest group, 2·19% vs 2·66% [RR 0·83, 95% CI 0·70, 0·98] in middle-aged adults, and 3·55% vs 4·41% [RR 0·81, 95% CI 0·72, 0·91] in senior adults. In contrast, vaccinated senior adults had a higher risk of IHD (14·87% vs 13·18% [RR 1·13, 95% CI 1·05, 1·21]) **(Supplementary Figure 1).**

## Discussion

In this large real-world retrospective cohort study, we examined risks for CRP outcomes in individuals with Long COVID using rigorously matched comparison groups and age- and sex-stratified analyses. Our novel findings demonstrate that Long COVID is associated with a persistent increase in multiple adverse CRP outcomes across all adult age groups, with a higher vulnerability in the young adults. Secondary analysis revealed that prior SARS-CoV-2 vaccination did not significantly reduce the risk for adverse CRP outcomes once Long COVID had developed. These results link the chronic health burden attributable to Long COVID and underscore the need for sustained surveillance and preventive strategies.

Our study design that compared outcomes in Long COVID patients directly to those who recovered without developing Long COVID, allowed us to isolate the cardiovascular risk attributable to Long COVID itself. We found significant increases in both AR and RR for major cardiovascular outcomes, with the greatest RR observed in the youngest adults and a larger effect was seen in young females. These findings suggest a possible disproportionate cardiovascular vulnerability in younger individuals, warranting mechanistic exploration of sex-based immune or vascular damage pathways distinct from traditional age-related mechanisms. Further studies are also needed to determine whether some of the difference might be related to other competing risks (e.g., all-cause mortality) or higher rates of other preventive measures (e.g., use of statins) in senior adults.

During the early pandemic months, the most common hematological outcomes observed was DVT or PE.^17–21^ Emerging studies have reported the existence of a prothrombotic state up to12 months post-acute COVID-19, suggesting persistence of long-term hypercoagulable state in these patients.^7,10,22–24^ However, these studies did not provide reference to patient’s Long COVID status, so the risk for thrombosis due to Long COVID per se remain elusive. Therefore, our findings in Long COVID patients demonstrating elevated risks for DVT and PE across all ages, and for IS in youngest and oldest adults, are novel. These findings also suggest a prolonged monitoring or consideration for thromboprophylaxis in the specific groups.

During acute COVID-19 infection, AKI was a frequent complication,^21,25–28^ yet prior studies of renal sequelae often included patients with preexisting kidney disease or prior AKI, complicating interpretation of Long COVID specific risk.^29–32^ Using a study design that excluded any patients with prior renal diseases and rigorously matching for renal risk factors, we observed an elevated risk for CKD at all ages and for AKI in younger adults. Our findings suggest that prior SARS-CoV-2 infection may serve as an independent risk factor for long-term renal dysfunction potentially through persistent tubular injury or immune-mediated mechanisms and call for prospective renal monitoring in Long COVID patients.

Notably, another important aspect of Long COVID is the progression to the long-term pulmonary complications.^33–39^. However, evidence for chronic respiratory outcomes beyond interstitial lung abnormalities remains limited.^40^ Our observation of a consistent increase in AR and RR for PF, PHT, CRF and CB across all ages, and the highest RR (10 times elevation) seen for PF in youngest male group suggest substantial susceptibility to long-term structural lung changes due to Long COVID. These outcomes may reflect persistent alveolar injury, microvascular thrombosis, or interactions with persistent cardiovascular dysfunction. Because we matched for asthma and COPD, our findings likely represent risks attributable directly to Long COVID rather than exacerbations of preexisting lung-conditions. Further investigation is needed to elucidate mechanisms distinguishing transient respiratory symptoms from progression to chronic fibrotic or other lung disease.

Consistent with prior studies,^41–43^ we also observed that females were more likely to develop Long COVID, yet absolute increases in CRP risks were similar between sexes. However, age-and sex-specific patterns in relative risk were prominent: young females had the highest relative risks for AF, IHD, HF, PE, AKI, and CKD, while young males showed disproportionately higher risks for PF and CB. These findings suggest that biological sex modulates vulnerability to organ-specific sequelae possibly mediated by hormonal influences, immune responses, or genetic factors. Understanding these mechanisms represents an important avenue for future research and may inform sex-specific diagnostic or preventive strategies.

Although vaccination reduces the risk of acute infection and lowers the likelihood of developing Long COVID,^44–47^ its protective effect once Long COVID is established remains uncertain. In our analysis, vaccination was associated with reduced risk of only HF among young adults and CRF across ages. The consistent reduction in CRF may indicate that vaccination may help limit persistent viral reservoirs, immune dysregulation, or microvascular injury in the lungs, the key mechanisms implicated in post-acute respiratory sequelae.^48–50^ Conversely, the association between vaccination and higher IHD risk in the senior adults is unexpected and may reflect residual confounding (e.g., baseline cardiovascular risk or healthcare-seeking behavior), or age-related differences in vascular vulnerability and immune responses. Overall, these findings indicate that vaccination alone may not sufficiently mitigate long-term complications in individuals who develop Long COVID, emphasizing the need for targeted therapeutic approaches.

### Limitations

Though our study identifies several novel findings, there are some limitations. *First*, using a de-identified database in TriNetX without direct review of patient records precludes the ability to verify the accuracy of patient documentation or obtain supplementary information. *Second,* patients may receive post-COVID care or vaccination outside the TriNetX network, leading to underreporting of outcomes and misclassification of vaccination status. *Third*, there is a possibility of coding variability across 109 health systems, as well as heterogeneity in long COVID definitions across sites. *Finally,* the TriNetX analytic tools used for this study did not permit patient-level, time-to-event analysis, so we were unable to assess risk differences in the cohort over time or to reliably measure potential mechanistic mediators (e.g., serological changes over time).

Despite these limitations, our study has several strengths including use of large real-world dataset, diversity of patients, use of rigorous cohort definitions, application of robust matching, age and sex-stratified risks, and reporting transparent methods. Thus, our findings have substantial long-term clinical implications.

## Conclusion

In this global, real-world study, Long COVID was independently associated with increased risk for adverse CRP outcomes. The relative risks were greatest in younger adults, with distinct sex-specific patterns. The absence of consistent protection from prior vaccination underscores the need for prolonged surveillance, mechanistic studies, and targeted preventive strategies. These findings have significant implications for healthcare systems, workforce productivity, and long-term care planning in patients with Long COVID.

## Acknowledgement

None

## Funding

This work was possible through fundings from National Institutes of Health AI162778, and HL168630, and Office of Research and Development and Department of Veterans Affairs 2I01BX007087 to S.D. TriNetX training and access provided by the Institute for Clinical and Translational Science at the University of Iowa, NIH CTSA program grant UM1TR004403.

## Author Contributions

SD and AP had full access to all the data in the study and take responsibility for the integrity of the data and the accuracy of the data analysis. A.P. participated in study design, collected data, and performed analysis. M.L.C., S.M.S., U.S. and A.P.C. provided scientific input on building cohorts, analysis and data interpretation. S.D. conceived the idea, directed the project, designed and interpreted the results, and co-wrote the manuscript. All authors assisted with the preparation and editing of the manuscript.

## Data availability

All data generated or analyzed are presented as part of this article or its supplemental material. The raw and analyzed data and materials related to this article but not included in it or in its supplemental material will be provided upon reasonable request. Any data and materials that can be shared will be released via a material transfer agreement. Data sharing statement

## Conflict of Interest Disclosures

Authors do not have any disclosures.

## Supplemental Material

Tables S1–S6

## Notes

### Competing Interest Statement

The authors have declared no competing interest.

### Clinical Trial

this is not a clinical trial

